# Global burden of foreign bodies and impact of COVID-19 pandemic: an analysis of 31 years of data and future forecasts

**DOI:** 10.64898/2026.04.29.26352072

**Authors:** Feiruzi Simayi, Kai Niu, Dongjie Li, Zhi Yu, Richeng Jiang

## Abstract

**Background:** Foreign bodies (FBs) can cause obstruction, infection, or injury, yet comprehensive global assessments remain limited. This study evaluated the burden of FBs from 1990–2021, projected trends to 2050, and identified high-risk populations.

**Methods:** Using Global Burden of Disease 2021 data, we estimated age-standardized incidence ratio (ASIR), death ratio (ASDR), and disability-adjusted life years (DALYs) by age, sex, and region. Temporal trends were assessed with estimated annual percentage change (EAPC) and Joinpoint regression; projections applied Bayesian age–period–cohort models; decomposition quantified the effects of aging, population, and epidemiological change.

**Results:** From 1990–2021, global ASIR declined from 660.75 to 561.16 per 100,000 (EAPC: –0.84), ASDR from 2.11 to 1.41 (–1.47), and DALYs from 145.14 to 77.87 (– 2.13). Males had consistently higher burden (2021: 725.96 versus 394.11 per 100,000 in females). Children under 5 and adults over 80 bore the highest risks, with intraocular FBs dominating incidence and pulmonary aspiration/airway FBs driving mortality. Western Europe had the highest ASIR, Andean Latin America the highest ASDR. Since 2019, the onset of the COVID-19 pandemic, intraocular FBs incidence has surged in East Asia, mainly China. Projections suggest ASIR will continue to rise through 2050, while ASDR and DALYs continue to decline, driven by global population growth (187.27%) and aging (46.82%) but offset by epidemiological improvements (−134.09%).

**Conclusions:** Despite long-term declines, FB incidence is rebounding, with marked disparities across sex, age, and region. Targeted interventions, including workplace safety, pediatric and geriatric care, and region-specific policies, are needed to mitigate risks and reduce inequalities.

## Introduction

Foreign bodies (FBs) are external objects or substances that enter the human body via ingestion, inhalation, insertion, or trauma, leading to mechanical obstruction, inflammation, or infection. Their clinical impact varies by anatomical site, material composition, and population demographics.

According to the Global Burden of Disease (GBD) 2021 classification, FBs are divided into three main categories based on anatomical location: Intraocular FBs (IOFBs), Pulmonary aspiration and FBs in airway (PAFBA), and FBs in other body parts which includes gastrointestinal tract, ear, nose, throat, and soft tissues. IOFBs penetrate the ocular wall, often injuring the vitreous and retina. IOFBs cause mechanical and chemical damage, resulting in visual loss, pain, and complications such as endophthalmitis and retinal detachment(1–4). Young and middle-aged men in industrial regions are particularly affected due to occupational hazards(1, 2). PAFBA primarily affect children under 5 years, who have immature swallowing reflexes and small airways. Common aspirated items include food, coins, and toys(5). In adults, aspiration risk increases among the elderly and patients with neuromuscular diseases such as stroke or Alzheimer’s disease(6, 7). Without timely intervention, aspiration may lead to hypoxic brain injury, pneumonia, airway stenosis, or even asphyxia. Gastrointestinal FBs are frequently ingested by children (coins, toys), and by adults (bones, button batteries)(8–10). High-risk groups include mentally ill individuals, edentulous adults, and prisoners(8, 11). While most FBs pass spontaneously, sharp objects can cause intestinal perforation, particularly in elderly patients due to delayed diagnosis(8, 12). FBs in the ear, nose, and throat are also common, especially in young children and adults with mental illness or after accidents. Clinical symptoms range from local irritation to airway obstruction(13–16). Technological advances such as magnetic conduction for IOFBs, flexible and fiberoptic bronchoscopy for airway FBs, and endoscopic removal for gastrointestinal FBs have improved treatment outcome(17–19). Emerging fields, including computer-assisted navigation, wearable sensors, and robotic-assisted surgery, show promise for enhancing prevention and treatment(20–23). Although the GBD 2021 study (24) provides the most authoritative and comprehensive overview of disease burden across 204 countries, it offers only brief descriptions for individual conditions, including FBs injuries. However, substantial knowledge gaps remain. Despite advancements in clinical practice, no study has quantified the joint contributions of population ageing, epidemiological change, and COVID-19-related behavioral shifts to FB burden across 204 countries. Furthermore, most existing studies lack comprehensive global and regional assessments disaggregated by age, sex, cause, and sociodemographic development. Tailored prevention and treatment strategies across populations and regions are urgently needed, along with stronger public education efforts. Building upon the GBD framework, this study presents the first systematic and in-depth analysis of the global, regional, and national burden of FBs from 1990 to 2021, projects trends to 2050, and identifies key priority groups and regions to inform targeted interventions and policy development.

## Methods

### Data Source

All data were retrieved from the GBD Results Tool of the Institute for Health Metrics and Evaluation (IHME). GBD 2021 data, collected via systematic reviews, government/international organization sources, primary data, and collaborator contributions, covers 371 diseases/injuries, 88 risk factors, 21 regions, and 204 countries/territories (1990–2021). Extracted indicators included age-standardized incidence rate (ASIR), age-standardized death rate (ASDR), and disability-adjusted life years (DALYs) stratified by 5-year age groups, sex, year, and region. ASIR/ASDR used GBD population standardization; DALYs combine years of life lost and lived with disability. All metrics include 95% uncertainty intervals (UI), with p < 0.05 for significance. Analyses and visualizations were done in R 4.4.2.

EAPC, calculated via least squares linear regression (ln(y) = α + βx + ε, where y = age-standardized rate, x = year), quantifies temporal trends: EAPC = (exp(β) − 1) × 100%. Trends were upward if 95% UI lower limit > 0, downward if upper limit < 0, and stable otherwise.

SDI, a composite of total fertility rate (under 25), average education (≥15), and lag-distributed income per capita (scaled 0–1), categorized 204 countries into five quintiles: low (0–0.4658), low-middle (0.4658–0.6188), middle (0.6188–0.7120), high-middle (0.7120–0.8103), high (0.8103–1.0).

### Disease definitions

In GBD 2021, FBs are defined as unintentional death or bodily damage from an extraneous material or substance being within the body, including the airway, lungs, nose, and eyes. ICD-9: 360.5-360.69, 374.86, 376.6, 709.4, 770.1-770.18, E911-E912.09, E913.0-E913.19, E913.8-E913.99, E914-E914.09, E915-E915.09; ICD-10: H02.81-H02.819, H44.6-H44.799, M60.2-M60.28, W44-W45, W45.3-W45.9, W75-W76.9, W78-W80.9, W83-W84.9.

## Statistical analysis

### Cross-country inequality analysis

Concentration Index (CI, range [-1,1]) quantified health inequalities by SDI. Negative CI indicates burden concentrated in low-SDI regions, positive in high-SDI; larger absolute values mean greater inequality. Countries were ranked by SDI, cumulative burden vs. population proportions calculated, Lorenz curves plotted, and CI with 95% UI derived via integration (R 4.4.2, two-tailed P < 0.05).

### Trend analysis

Compound line graphs visualized trends in ASIR, ASDR, and DALYs related to FBs. Data were stratified according to GBD standards, including 5-year age groups, sex (male/female), SDI regions (low/low-middle/middle/high-middle/high), and time (1990–2021). Data analysis and visualization were performed using R software to intuitively display temporal trends and inter-group differences in FBs disease burden across different dimensions.

### Joinpoint regression analysis

Joinpoint Regression Software (5.4.0) identified inflection points via segmented log-linear regression. Optimal joinpoints (0–5) were selected using Bayesian information criterion and Monte Carlo permutation tests (5000 iterations). Annual percentage change (APC) and 95% CIs were calculated; average APC weighted by segment time span. Trends were significant if 95% CI excluded 0.

### Decomposition analysis

This study adopted the Das Gupta decomposition method to dissect changes in ASDR of FBs from 1990 to 2021, isolating the independent contributions of aging, population growth, and epidemiological changes. Unlike traditional linear regression that focuses on variable correlations, this method enables precise quantification of each factor’s unique impact on overall burden shifts. The total burden for each region was computed using the formula: D_total(y) = Σ(ai,y × Py × ei,y), where ai,y represents the proportion of the population in the i-th age group (among 20 age groups) in year y, Py is the total population in year y, and ei,y denotes the age-specific ASDR for the i-th age group in year y. By holding other variables constant, the distinct effect of each factor on ASDR changes was measured—positive values indicate the factor increases burden, while negative values reflect a burden-reducing effect.

### BAPC model projection

The Bayesian age–period–cohort (BAPC) model was selected for future burden forecasting, tailored to handle the complex, high-dimensional, and sparse datasets characteristic of large-scale epidemiological studies like GBD 2021. Rooted in the generalized linear model framework and adapted to a Bayesian setting, the model dynamically integrates age-, period-, and cohort-specific effects. These effects are assumed to evolve over time and are smoothed through second-order random walks, enhancing the precision of posterior probability predictions. A key advantage of the BAPC model is its use of integrated nested Laplace approximation for estimating marginal posterior distributions, which avoids issues like poor mixing and convergence challenges associated with Markov chain Monte Carlo techniques while maintaining high computational efficiency. Widely validated in epidemiological research—especially for age-structured population data and complex cohort effects—the model was implemented via the "BAPC" R package. It utilized GBD 2021 data and IHME population projections to forecast the global burden of FBs and their subtypes, capturing complex interactions between age, period, and cohort effects for nuanced future predictions.

## Results

### Global and Regional Burden of FBs

From 1990 to 2021, the global ASIR of FBs decreased from 660.75 (95% UI: 522.55 to 868.74) to 561.16 (95% UI: 441.12 to 728.48) per 100,000 population, with an EAPC of −0.84 (95% UI: −1.06 to −0.62). Males showed significantly higher ASIR than females (725.96 [95% UI: 568.26 to 964.09] vs 394.11 [95% UI: 316.05 to 493.99] per 100,000 in 2021), though with a more pronounced annual decline (EAPC: −0.92 [95% UI: −1.18 to −0.65] vs −0.69 [95% UI: −0.84 to −0.53]) (**Table 1**).

**Table 1.**
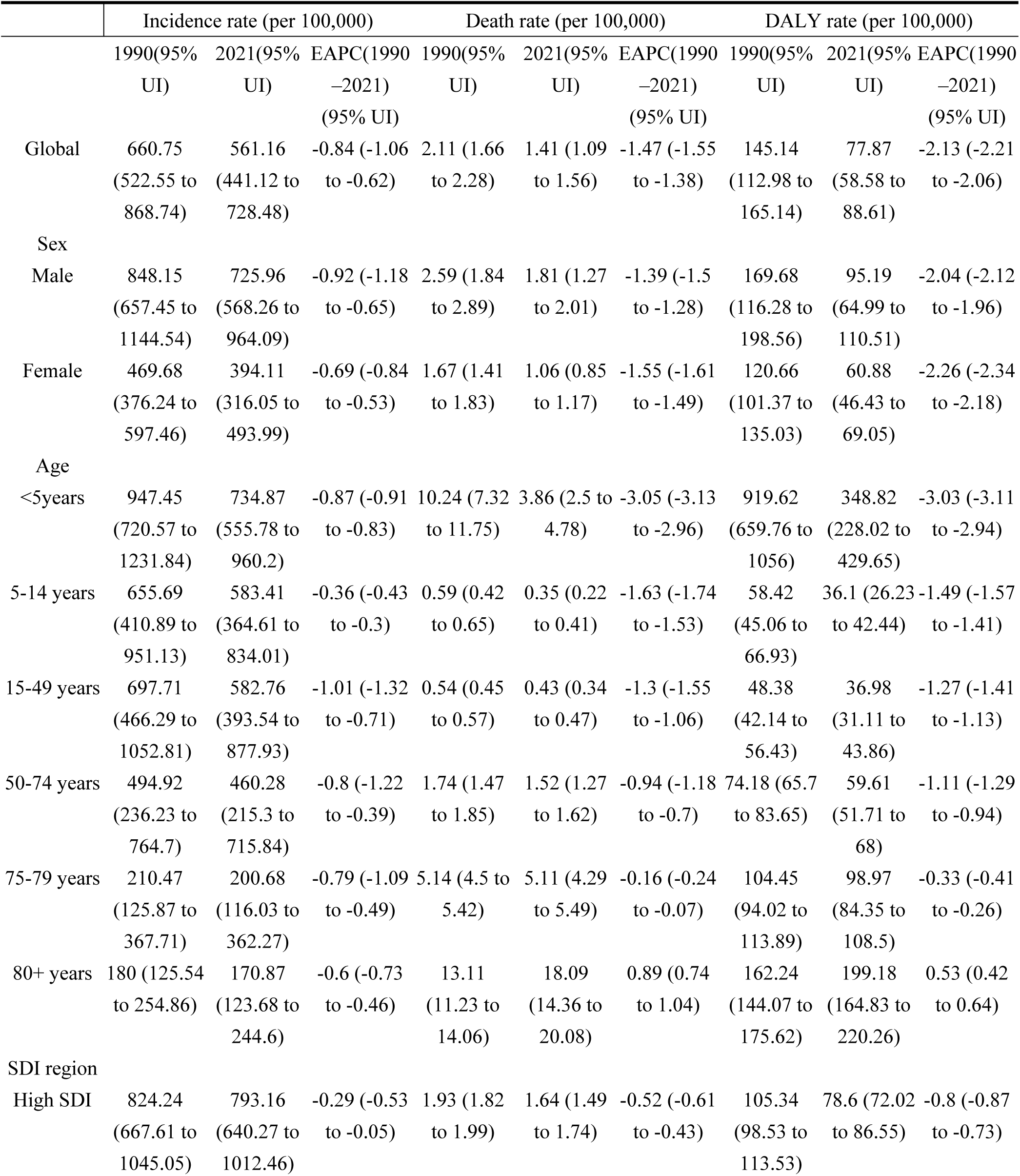
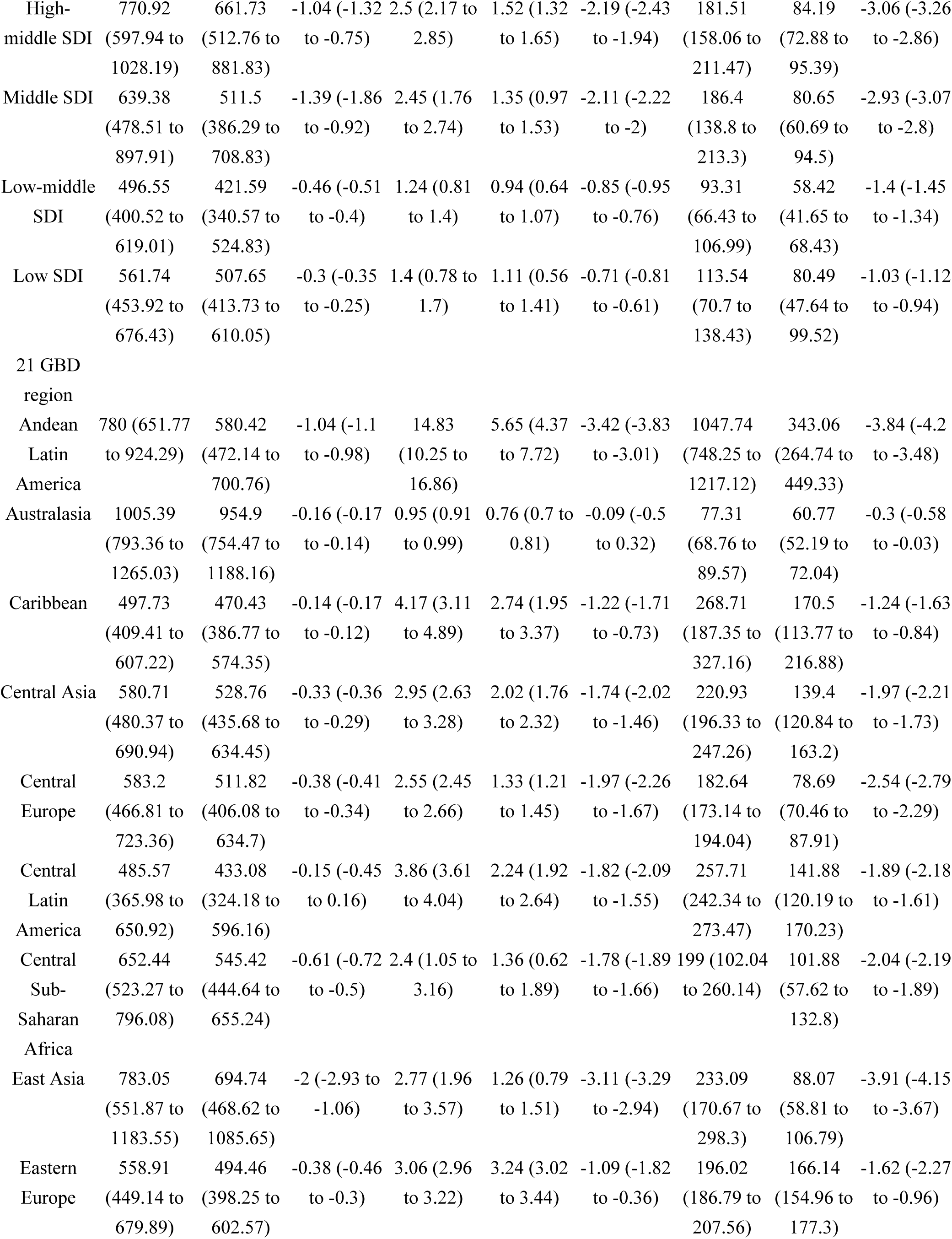
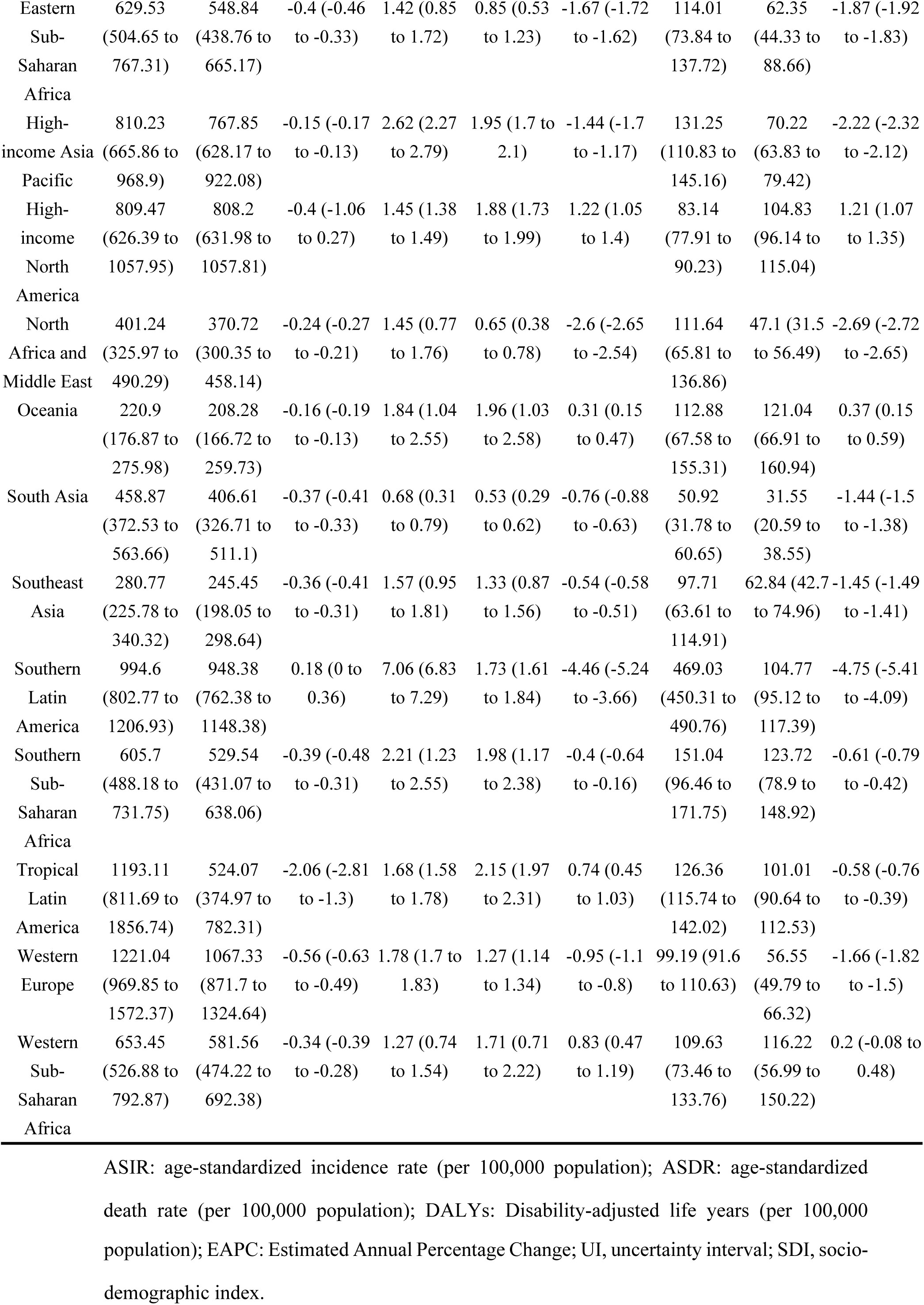
The ASIR, ASDR, and DALYs related to foreign bodies in 1990 and 2021, along with their changes from 1990 to 2021, analyzed globally, by sex, age group, SDI regions, and 21 GBD regions.

Similar patterns emerged for mortality, with the ASDR decreasing from 2.11 (95% UI: 1.66 to 2.28) in 1990 to 1.41 (95% UI: 1.09 to 1.56) in 2021, with an EAPC of −1.47 (95% UI: −1.55 to −1.38), and males again showing higher ASDR than females (1.81 [95% UI: 1.27 to 2.01] vs 1.06 [95% UI: 0.85 to 1.17]) (**Table 1**). In addition, DALYs caused by FBs declined from 145.14 (95% UI: 112.98 to 165.14) to 77.87 (95% UI: 58.58 to 88.61), with an EAPC of −2.13 (95% UI: −2.21 to −2.06), maintaining higher male burden (95.19 [95% UI: 64.99 to 110.51] vs 60.88 [95% UI: 46.43 to 69.05]) (**Table 1**).

Among the 21 GBD regions, Western Europe had the highest ASIR (1067.33 [95% UI: 871.7 to 1324.64]) in 2021, followed by Australasia [954.9 (95% UI: 754.47 to 1188.16)] and Southern Latin America [948.38 (95% UI: 762.38 to 1148.38)], whereas Oceania showed the lowest (208.28 [95% UI: 166.72 to 259.73]). Southern Latin America was the only region with increasing ASIR (EAPC: 0.18 [95% UI: 0 to 0.36]), contrasting with steep declines in East Asia (EAPC: −2.29 [95% UI: −2.93 to −1.06]) and Tropical Latin America (EAPC: −2.06 [95% UI: −2.81 to −1.30]) (**Table 1**).

The ASDR was highest in Andean Latin America (5.65 [95% UI: 4.37 to 7.72]) and Eastern Europe (3.24 [95% UI: 3.02 to 3.44]), with increases noted in high-income North America (EAPC: 1.22 [95% UI: 1.05 to 1.4]) and Western sub-Saharan Africa (EAPC: 0.83 [95% UI: 0.47 to 1.19]). DALYs peaked in Andean Latin America (343.06 [95% UI: 264.74 to 449.33]) and rose slightly in high-income North America (EAPC: 1.21 [95% UI: 1.07 to 1.35]), whereas declined markedly in Southern Latin America (EAPC: −4.75 [95% UI: −5.41 to −4.09]) and Andean Latin America (EAPC: −3.84 [95% UI: −4.2 to −3.48]) (**Table 1**).

### Burden across countries and SDI quintiles

At the national level, Italy, Finland, New Zealand, and Belgium reported the highest ASIR in 2021, however, Italy showed the most significant decrease from 1990 to 2021. Although most countries experienced a decline in ASIRs from 1990 to 2021, several exceptions were noted, including Chile, Brunei Darussalam, Cuba, and 11 other territories. Notable annual reductions in ASIR were observed in Brazil (EAPC: −2.11 [95% UI: −2.88 to −1.33]) and China (EAPC: −2.03 [95% UI: −2.98 to −1.08]) (**Figure 1A, B**, Supplementary Table S1).

**Figure 1.**
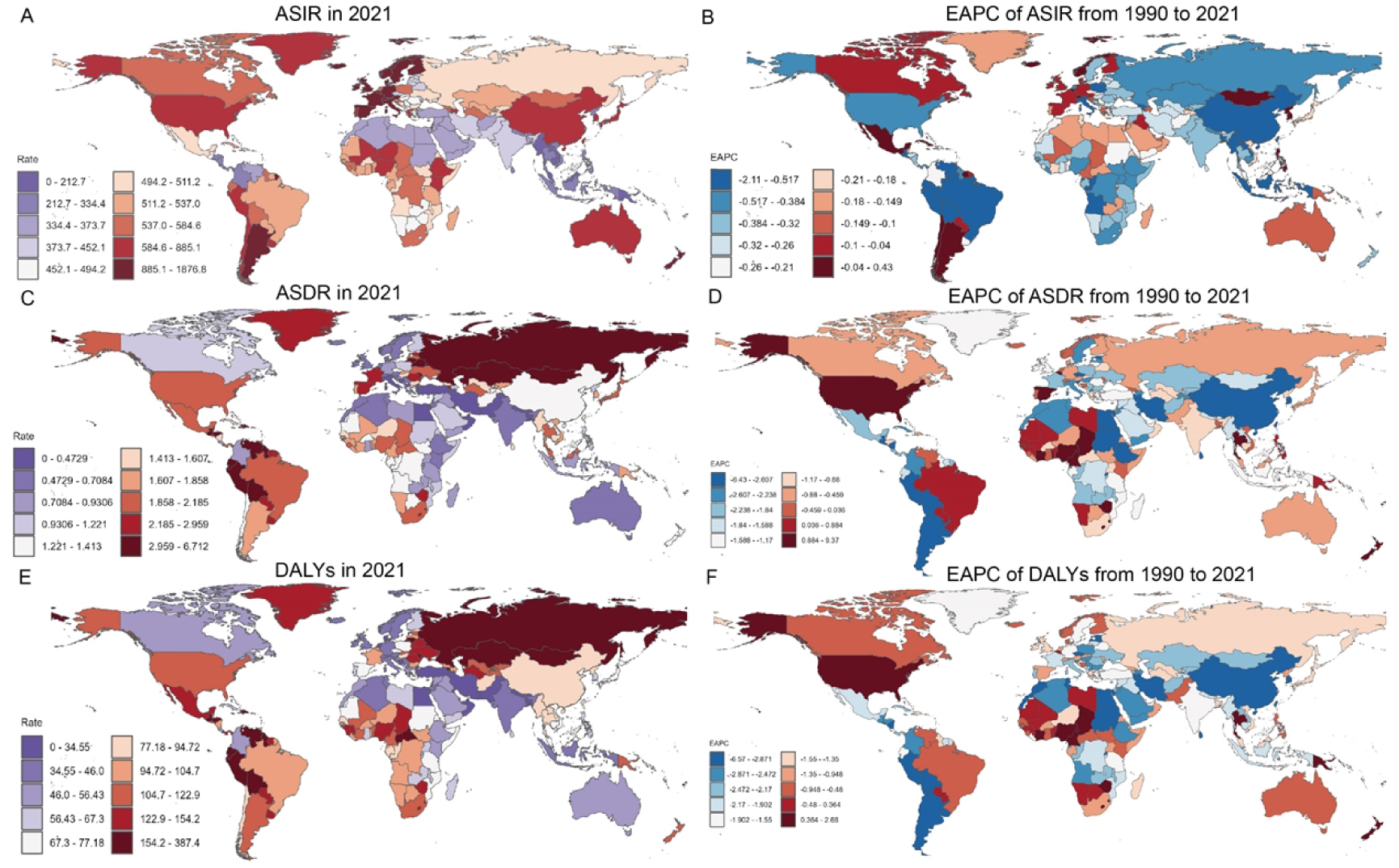
Global burden and trends of foreign bodies across 204 countries and territories. (A) ASIR in 2021; (B) EAPC of ASIR from 1990 to 2021; (C) ASDR in 2021; (D) EAPC of ASDR from 1990 to 2021; (E) DALYs in 2021; (F) EAPC of DALYs from 1990 to 2021. ASIR: age-standardized incidence rate (per 100,000 population); ASDR: age-standardized death rate (per 100,000 population); DALYs: Disability-adjusted life years (per 100,000 population); EAPC: Estimated Annual Percentage Change.

Honduras, Peru, and Bolivia recorded the highest ASDR. Notable declines in ASDRs were seen in Chile (EAPC: −6.43 [95% UI: −7.89 to −4.95]) and Egypt (EAPC: −5.67 [95% UI: −6.17 to −5.18]), though Cabo Verde exhibited a striking increase (EAPC: 9.37 [95% UI: 6.00 to 12.86]) (**Figure 1C, D**).The highest disease burden measured by DALYs concentrated in Bolivia and Peru, with notable reductions in Chile (EAPC: - 6.57 [95% UI: −8.04 to −5.07]) and Uruguay (EAPC: −5.21 [95% UI: −5.72 to −4.70]) **(Figure 1E, F**, Supplementary Table S1).

By SDI quintile, ASIRs were higher in high-SDI and high-middle-SDI regions, particularly in Western European countries, where ASIRs demonstrated an increasing trend with rising SDI levels. Conversely, ASDRs and DALYs were concentrated in middle-SDI regions, followed by a slight decline in these metrics as SDI increased (**Figure 2**).

**Figure 2.**
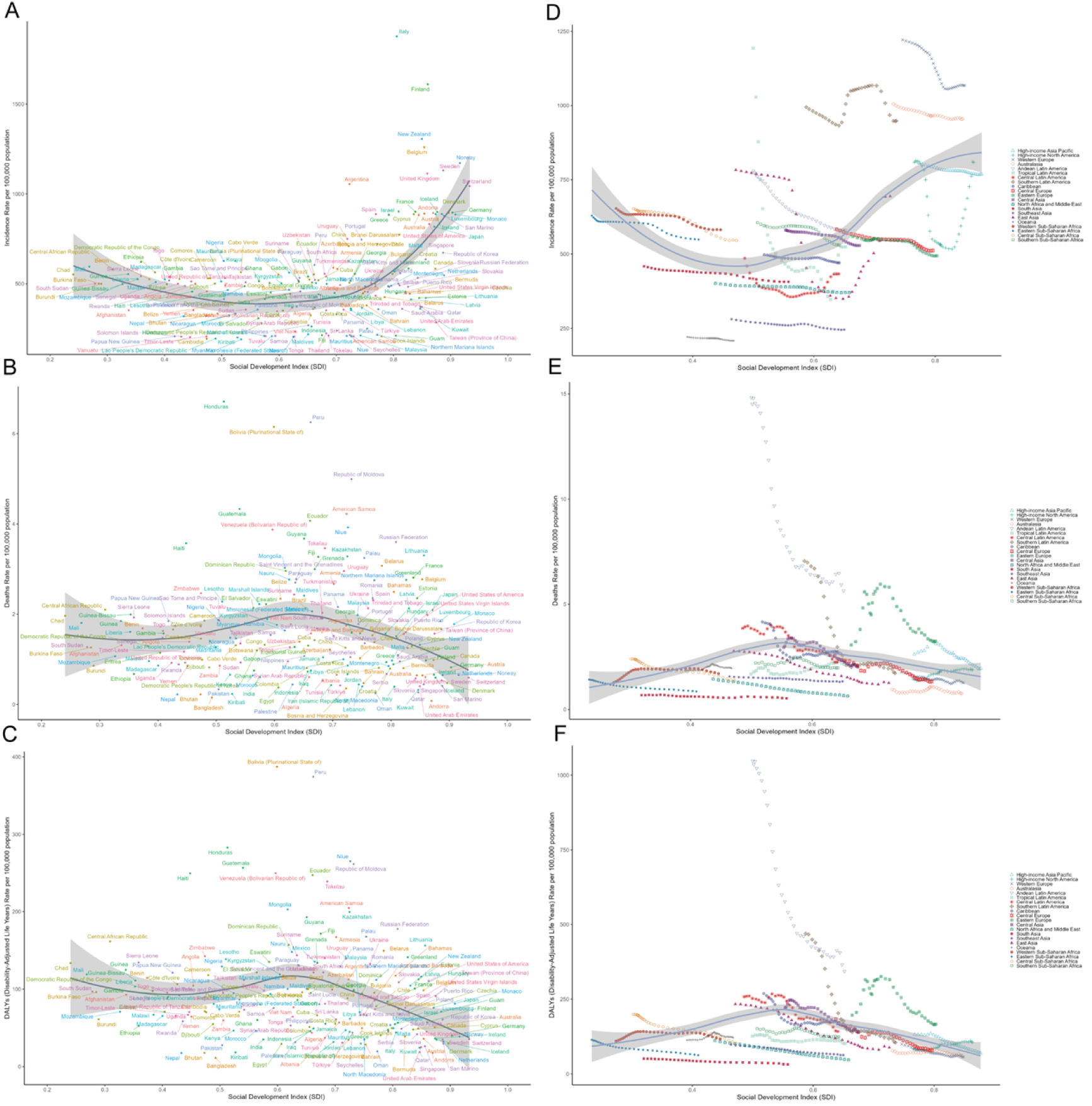
The ASIR,ASDR and DALYs for foreign bodies in 204 countries and territories. (A)ASIR; (B)ASDR; (C)DALYs for 204 countries and (D)ASIR; (E)ASDR; (F)DALYs for GBD regions by Socio-demographic Index, 1990–2021. Expected values based on the Socio-demographic Index and disease rates in all locations are shown as the black line. This black line represents the expected age-standardized rates for each value of the Socio-demographic Index, and is based on a Gaussian process regression of results from all Global Burden of Disease locations over the entire 1990–2021 estimation period. ASIR: age-standardized incidence rate(per 100,000 population); ASDR: age-standardized death rate(per 100,000 population);DALY=disability-adjusted life-year(per 100,000 population).

### Age-, sex-, time-specific burden

From 1990 to 2021, the ASIRs of FBs declined in both sexes. However, a more pronounced age disparity was observed in males, with incidence remaining heavily concentrated among younger age groups, particularly the age 35-44 and children under five years old (**Supplementary Figure S1A**). In contrast, no significant sex-based differences were found in ASDR and DALYs. Both exhibited a higher disease burden among children under five years old and the elderly over 80 years old (**Supplementary Figure S1B, C**).

When stratified by SDI, children under five consistently showed higher ASIR, ASDR, and DALYs than age-standardized values, though with a declining trend. Meanwhile, ASDR and DALYs in adults aged 80 years and above increased over time, surpassing those in children by 2021, especially in high-SDI regions, where the DALY gap between the two groups nearly disappeared (**Figure 3**).

**Figure 3.**
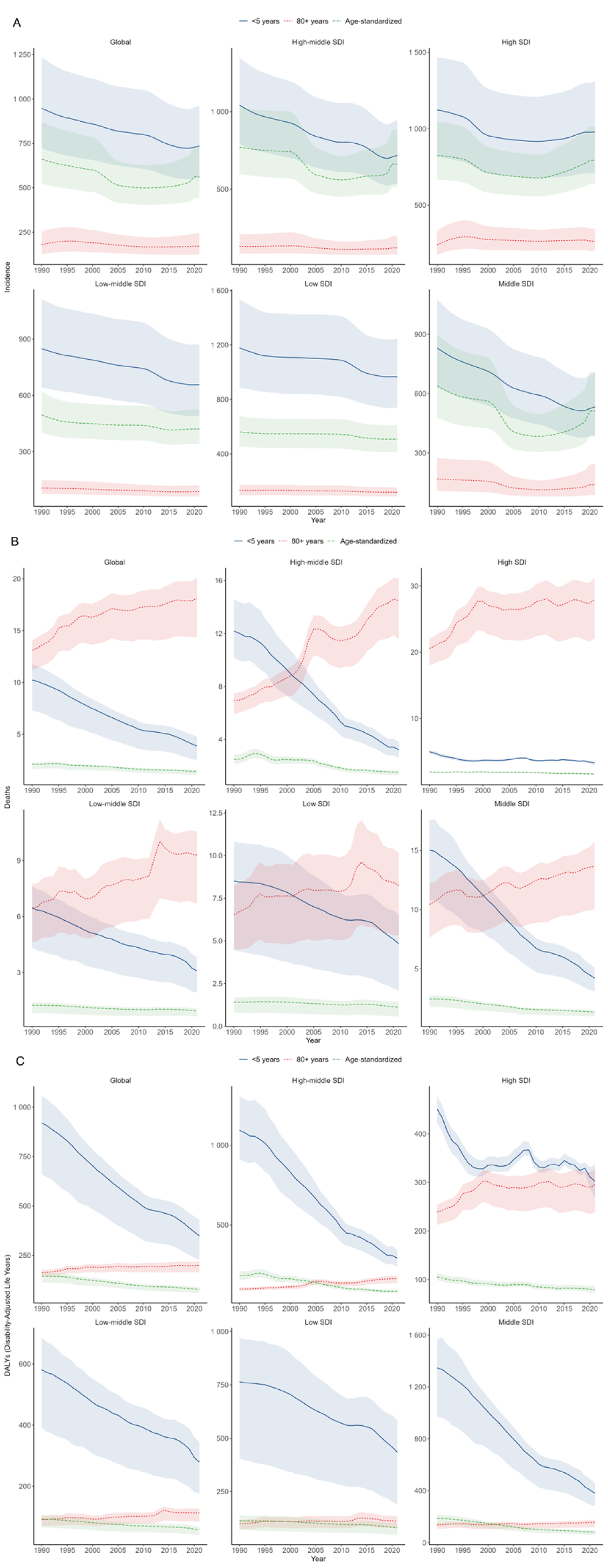
Temporal trends of ASIR and ASDR of foreign bodies globally and in different SDI regions from 1990 to 2021, by age group (<5years, 80+years, Age-standardized). The changes in the (A) incidence rates; (B) death rates; (C) DALYs rate of children (<5 years), the elderly (80+years), and age-standardized incidence rates globally and across different SDI regions from 1990 to 2021. DALYs: Disability-adjusted life years

### Burden by foreign body subtype

Among FB types, IOFBs had the highest ASIRs, especially in individuals aged 30–55. FBs in other body parts and PAFBA were more common in children under five and adults over 80. PAFBA was particularly elevated in children (**Supplementary Figure S2A**).

In terms of ASDR, FBs in other body parts exhibited relatively uniform mortality rates across all age groups, while PAFBA began to increase markedly after the age of 80. IOFBs, as expected, were associated with no mortality and thus did not contribute to ASDR (**Supplementary Figure S2B**). For DALYs, IOFBs and FBs in other body parts did not display significant age-related disparities, whereas PAFBA consistently imposed a considerable disease burden in children under five and adults over 80 years (**Supplementary Figure S2C**).

To further elucidate regional differences, a heatmap analysis was conducted across 21 GBD regions. IOFBs were more prevalent in high-income regions, Western Europe, and East Asia. FBs in other body parts were concentrated in Australasia and Southern Latin America. PAFBA had globally low incidence but contributed substantially to DALYs in Latin America, Eastern Europe, Central Asia, Oceania, and sub-Saharan Africa (**Supplementary Figure S3**).

### Joinpoint regression and future projections

Joinpoint regression revealed a biphasic global ASIR trend, with modest declines during 1990-2001 (APC: −0.96) and 2005-2010 (APC: −0.53), a sharp drop in 2001-2005 (APC: −3.57), and a rising trend thereafter: 2010-2015 (APC: 0.27), 2015-2018 (APC: 1.10), and 2018-2021 (APC: 2.81), indicating accelerated incidence in recent years (**Figure 4A**). ASDR declined steadily except for a brief increase during 1990-1994 (APC: 1.15) (**Figure 4B**). DALYs showed continuous reductions, with the fastest declines during 1994-1997 (APC: −2.82) and 2004-2010 (APC: −2.78) (**Figure 4C**).

**Figure 4.**
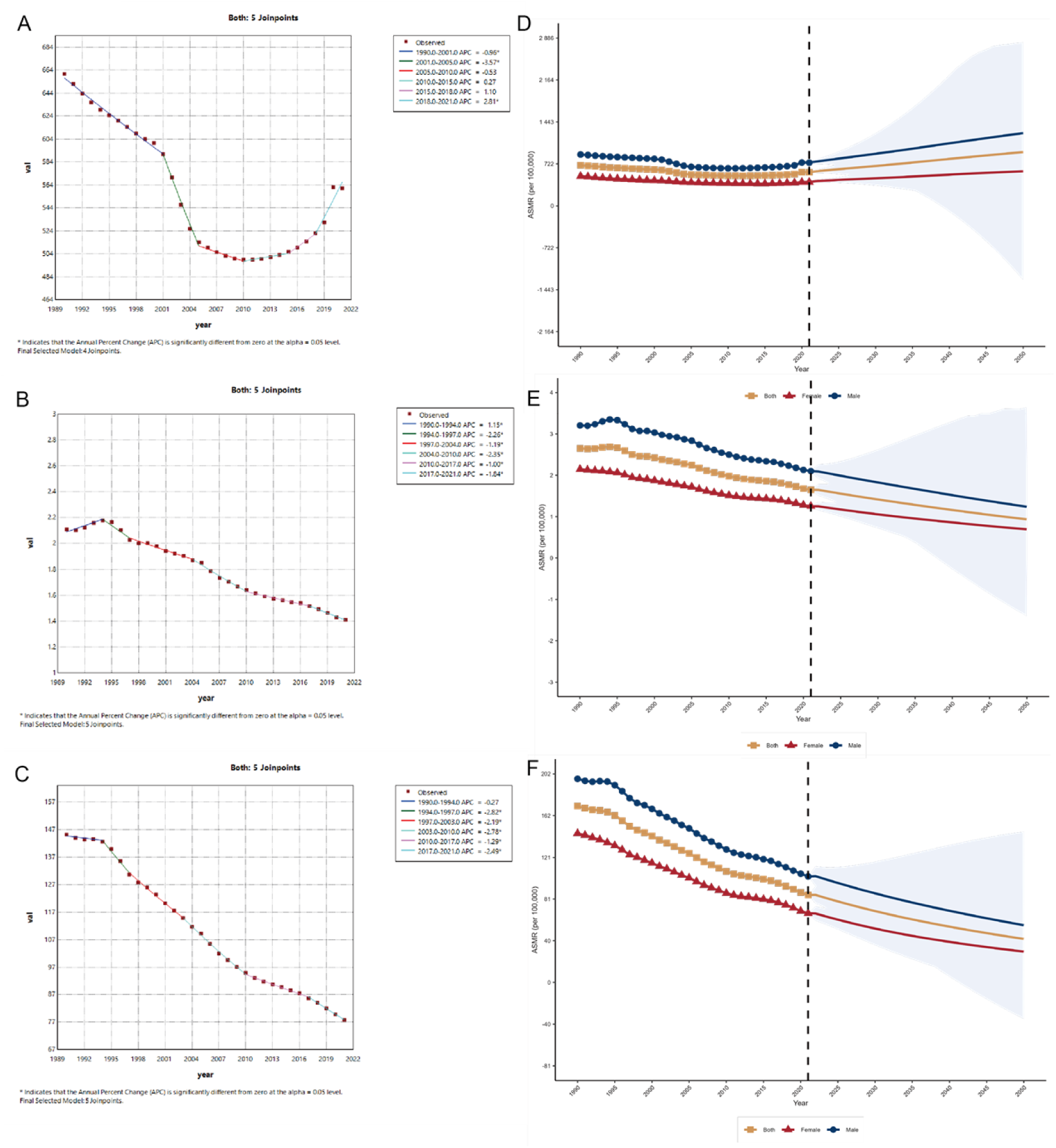
Joinpoint regression analysis results: Annual percent change (APC) and trends in global burden of foreign bodies from 1990 to 2021. Joinpoint regression analysis of foreign bodies from 1990 to 2021. (A) ASIR; (B) ASDR; (C) DALYs rate. Red squares represent actual observed values, and colored line segments denote the fitted trend lines for each time period identified by the Joinpoint model. Values labeled next to each line segment are the APCs; an asterisk (*) indicates that the APC is statistically significant at the p<0.05 level. Estimated trends in the global burden of foreign bodies by sex, with 95% uncertainty intervals, 2021–2050. (D) ASIR; (E) ASDR; (F) DALYs rate. Results after the dashed line represent BAPC-based projections; yellow represents the total population, blue represents males, and red represents females, with the blue shaded area corresponding to the 95% Bayesian credible interval. ASIR: age-standardized incidence rate(per 100,000 population); ASDR: age-standardized death rate (per 100,000 population); DALYs: Disability-adjusted life years (per 100,000 population).

BAPC models project a continued rise in the ASIR through 2050, while ASDR and DALYs are expected to decline, reflecting advances in clinical care (**Figure 4D, E, F**). The male population consistently shows higher burden and is projected to remain at greater risk.

### Subtype-specific surge post-2019 (post-COVID-19)

Given the sharp rise in ASIR after 2019 (**Supplementary Figure S4A**), we further analyzed the three FB subtypes during 2019-2021 specifically. Notably, IOFBs exhibited increasing ASIRs across most regions, with significant rises in East Asia and high-income North America during 2020 and 2021 (**Supplementary Figure S4A**). Additionally, in East Asia, all three FB types showed rising incidence, whereas other regions exhibited stable trends (**Supplementary Figure S4A, B, C**). Furthermore, Country-level analysis revealed that the rise in IOFBs ASIR in East Asia was predominantly driven by China, with notable contributions also from the United States **Supplementary Figure S4D**). The growth in PAFBA and FBs in other body parts was also particularly attributed to China, whereas declines were more pronounced in the United States and Brazil (**Supplementary Figure S4E, F**).

### Inequality and Decomposition Analysis

Between 1990 and 2021, the CI for FB-related DALYs fell from 0.10 to-0.06, indicating a shift toward a more equitable global distribution. Initially, FB burden was concentrated in higher SDI regions, but the negative CI in 2021 reflects a relative burden shift to lower SDI populations (**Figure 5A**).

**Figure 5.**
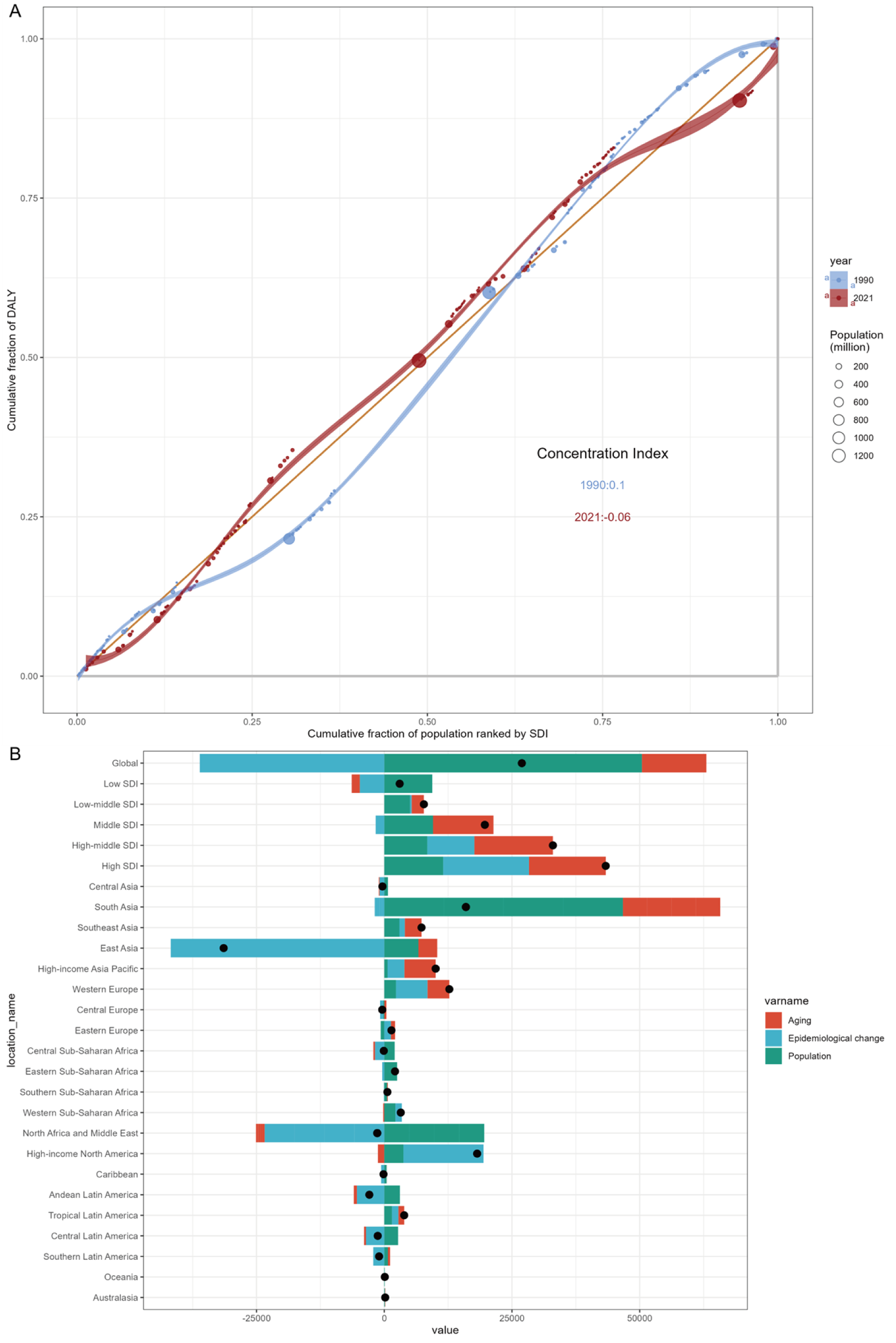
SDI-Related DALY Inequalities and ASDR Determinants for Foreign Bodies, 1990–2021. (A) Relative SDI-related inequalities in DALYs in 204 countries and territories, 1990–2021; (B) Population-level determinant changes of ASDR in aging, population growth, and epidemiological changes for foreign bodies globally and in various regions from 1990 to 2021. Black dots represent the total change contributed by all three components. A positive value for each component indicates a corresponding positive contribution in ASDR, and a negative value indicates a corresponding negative contribution in ASDR. ASDR: age-standardized death rate (per 100,000 population); DALYs: Disability-adjusted life years (per 100,000 population).

Decomposition analysis revealed that population growth (187.27%) and aging (46.82%) were the main drivers of burden increases. Epidemiological improvements offset part of this rise, contributing a 134.09% reduction (**Supplementary Table S2**). Aging had the most significant impact in high-, high-middle-, and middle-SDI regions, especially in South and East Asia. Epidemiological improvements, likely due to better healthcare access and awareness, were especially influential in East Asia (**Figure 5B**).

## Discussion

This study provides the most comprehensive assessment to date of the global, regional, and national burden of FBs from 1990 to 2021, with projections to 2050 using GBD 2021 data. Between 1990 and 2021, global ASIR, ASDR, and DALYs due to FBs generally declined, reflecting improvements in public health, workplace safety, and healthcare accessibility. However, the post-2010 rebound in ASIR, particularly after 2018, is concerning. This trend may be driven by industrial expansion in emerging economies and indicates the urgent need for strengthened prevention strategies. While ASDR and DALYs are projected to decline further, likely due to advances in emergency care and medical technologies, the continued rise in ASIR demands attention.

Gender disparities in FB incidence are closely linked to occupational exposure, behavioral traits, and population-specific habits. Males are overrepresented in high-risk occupations such as construction, welding, carpentry, mining, and agriculture, contributing to the elevated incidence of IOFBs(25, 26). Additionally, males are more likely to experience weapon-related eye injuries during military service. For airway FBs, children and males are more prone to inhalation, related to males being more active, risk-prone, and more likely to engage in outdoor or competitive activities. In studies of genitourinary FBs, 24.8% of male cases required hospitalization, far higher than 2.1% of female cases, due to males more frequently having urethral or bladder FBs that often require endoscopic or surgical intervention(27).

Children and the elderly are most vulnerable to severe FB outcomes. However, their epidemiological trends show distinct differences: the incidence among children has been on the decline, while that among the elderly is rising. Notably, in high SDI regions, the gap in DALYs between the two groups is narrowing. In children, anatomical and behavioral factors such as immature dentition, impaired airway protection, and developmental disorders increase risk(28–30). In the elderly, burden is driven by delayed retirement, poor vision, impaired swallowing, neurological conditions, and medication effects(26, 28).

Among FB types, IOFB had the highest ASIR, while PAFBA had the highest ASDR and DALYs, especially in those under five and over 80. High- and high-middle-SDI regions (e.g., Western Europe) had the highest ASIRs, increasing with SDI, while ASDR and DALYs were concentrated in middle-SDI regions (e.g., Andean Latin America), declining with SDI. The decline in East Asia, particularly China, reflects successful policy interventions such as the 2009 workplace safety regulations and bans on fireworks.

Notably, FB incidence rose again after 2019, especially IOFBs in East Asia, primarily China. The COVID-19 pandemic contributed to this trend by increasing children’s exposure to small household objects, reducing caregiver supervision, and delaying access to medical care. Pediatric hospital reports from China documented sharp increases in coin and battery ingestion during COVID-19 pandemic. Factors such as poor mask quality, psychological stress, and treatment delays further compounded the risk.

Regional disparities in FB burden are closely tied to socioeconomic development. From 1990 to 2021, the distribution of disease burden became more equitable, with reduced burden on socially disadvantaged groups. High-income countries benefit from healthcare infrastructure, early detection, and regulatory oversight. In contrast, low- and middle-income countries, such as those in sub-Saharan Africa, face rising FB burdens due to occupational hazards, limited safety equipment, and ongoing conflict(26, 31). Industrial transformation, particularly in mining and agriculture, has also increased exposure in these regions.

Multiple studies support the effectiveness of preventive strategies against FBs. Evidence-based policies, including vehicle safety features and fireworks bans have significantly reduced related eye and airway injuries, while personal protective equipment can prevent over 90% of work-related IOFBs in occupational and sports settings(32). However, low compliance due to limited awareness and institutional support underscores the need for stronger public education and policy enforcement. Tailored interventions remain essential: for children, caregiver education and product safety; for the elderly, dental care and mental health management; for workplaces, enforcement of personal protective equipment and safety training. Advances in diagnostics, including computed tomography, ultrasound, and radiography facilitate FB detection(33). In addition, minimally invasive endoscopy reduces complications and

AI-driven imaging tools show promise for FB identification and MRI safety enhancement(34). Mobile health technologies further improve surveillance and management by enabling real-time data collection, decision support, and cost reduction(35). Establishing a cross-national, real-time FB registry could standardize data reporting, foster international collaboration, and enable dynamic preventive strategies, ultimately reducing the global FB burden.

This study highlights the urgency of addressing the renewed rise in ASIR after 2010, especially the surge post-2019, and calls for tailored interventions targeting high-risk populations and regions. Coordinated international efforts, policy exchange, and expanded use of protective strategies are critical for achieving equitable reductions in FB burden worldwide.

## Conclusion

Overall, global FB-related ASIR declined initially but rebounded after 2010, accelerating post-2018 and projected to rise through 2050, while ASDR and DALYs continued to decline. Disparities remain across regions, age, and sex, with children and the elderly bearing the highest burden and IOFBs and PAFBA as major contributors. The post-2019 surge in IOFBs in East Asia, especially China, highlights the need for targeted prevention and further investigation.

## Data Availability

No data were generated in this study. All analyzed data were obtained from the Global Burden of Disease (GBD) 2021 database, which is publicly available through the Global Health Data Exchange (GHDx) platform at https://ghdx.healthdata.org/ and the GBD Results Tool at https://vizhub.healthdata.org/gbd-results/.

## List of abbreviations

APC: Annual percentage change
ASIR: Age-standardized incidence rate
ASDR: Age-standardized death rate
BAPC: Bayesian age-period-cohort
CI: Confidence interval
COVID-19: Corona Virus Disease 2019
DALYs: Disability-adjusted life years
EAPC: Estimated annual percentage change
FBs: Foreign bodies
GBD: Global Burden of Disease
IHME: Institute for Health Metrics and Evaluation
IOFBs: Intraocular foreign bodies
PAFBA: Pulmonary aspiration and foreign bodies in the airway
SDI: Socio-demographic Index
UI: Uncertainty interval

## Ethics approval and consent to participate

This study uses data from the GBD Study 2021, which has been reviewed and approved by the Institutional Review Board of the University of Washington, with a waiver of informed consent (https://www.healthdata.org/researchanalysis/gbd).

## Consent for publication

Not applicable.

## Availability of data and materials

The data that support the findings of this study are openly available in the GBD 2021 database, which is publicly accessible via the Global Health Data Exchange (GHDx) platform (https://ghdx.healthdata.org/). Detailed methods and datasets can be retrieved from the GBD Results Tool (https://vizhub.healthdata.org/gbdresults/). Further inquiries should be directed to the corresponding author.

## Author contributions

RJ initiated and designed the study. FS contributed to the conceptualization, methodology, data analysis, and writing the initial draft of the manuscript. KN contributed to data validation and reviewing manuscript. DL, ZY contributed to manuscript review and editing. All authors read and approved the final version.

## Acknowledgments

We would like to express our sincere gratitude to the Institute for Health Metrics and Evaluation at the University of Washington, the GBD collaborators, and all staff members who contributed data to this research. It should be noted that the opinions put forward in this paper are exclusively those of the authors and do not reflect the official stances of their associated institutions.

## References

1. Liang Y, Liang S, Liu X, Liu D, Duan J. Intraocular Foreign Bodies: Clinical Characteristics and Factors Affecting Visual Outcome. J Ophthalmol. 2021;2021:9933403.

2. Liu CCH, Tong JMK, Li PSH, Li KKW. Epidemiology and clinical outcome of intraocular foreign bodies in Hong Kong: a 13-year review. International Ophthalmology. 2017;37(1):55–61.

3. Lewin-Smith MR, Strausborger SL, Jenkins HM, Merezhinskaya N, Latkany PA, Mazzoli RA, et al. The Joint Pathology Center/Vision Center of Excellence Approach to Analyzing Intra-Ocular “Foreign Bodies”. Military Medicine. 2019;184(Supplement_1):565–70.

4. Loporchio D, Mukkamala L, Gorukanti K, Zarbin M, Langer P, Bhagat N. Intraocular foreign bodies: A review. Survey of Ophthalmology. 2016;61(5):582–96.

5. Salih AM, Alfaki M, Alam-Elhuda DM. Airway foreign bodies: A critical review for a common pediatric emergency. World J Emerg Med. 2016;7(1):5–12.

6. Hewlett JC, Rickman OB, Lentz RJ, Prakash UB, Maldonado F. Foreign body aspiration in adult airways: therapeutic approach. J Thorac Dis. 2017;9(9):3398–409.

7. Hong P-Y, Wang L, Du Y-P, Wang M, Chen Y-Y, Huang M-H, et al. Clinical characteristics and removal approaches of tracheal and bronchial foreign bodies in elders. Scientific Reports. 2024;14(1):9493.

8. Erbil B, Karaca MA, Aslaner MA, Ibrahimov Z, Kunt MM, Akpinar E, et al. Emergency admissions due to swallowed foreign bodies in adults. World J Gastroenterol. 2013;19(38):6447–52.

9. Kramer RE, Lerner DG, Lin T, Manfredi M, Shah M, Stephen TC, et al. Management of Ingested Foreign Bodies in Children. Journal of Pediatric Gastroenterology and Nutrition. 2015;60(4):562–74.

10. Smith MT, Wong RK. Foreign bodies. Gastrointest Endosc Clin N Am. 2007;17(2):361–82, vii.

11. Hsieh A, Hsiehchen D, Layne S, Ginsberg GG, Keo T. Trends and clinical features of intentional and accidental adult foreign body ingestions in the United States, 2000 to 2017. Gastrointest Endosc. 2020;91(2):350–7.e1.

12. Zhang Hong YS, Wang Yanhong, Zhang Yaowei, Luan Zhaohui, Lin Hui, Bai Jianying, Yang Shiming, Fan Chaoqiang. Risk factors and treatment strategies of digestive tract foreign body perforation. Journal of Army Medical University. 2022;44(4): 379–84.

13. Abraham ZS, Kahinga AA, Khamis KO, Liyombo E. Clinical spectrum of ear, nose and throat foreign bodies at a tertiary hospital: a cross-sectional study. Ann Med Surg (Lond). 2023;85(7):3403–8.

14. Kekre M, Chakravarty S, Agarwal R. Foreign Bodies in Ear, Nose, Throat and Maxillofacial Region: A Study on Their Clinical Profile and Complications. Indian J Otolaryngol Head Neck Surg. 2022;74(Suppl 3):4483–94.

15. Heim SW, Maughan KL. Foreign bodies in the ear, nose, and throat. Am Fam Physician. 2007;76(8):1185–9.

16. Grigg S, Grigg C. Removal of ear, nose and throat foreign bodies: A review. Australian Journal for General Practitioners. 2018;47:682–5.

17. Wang Y, Chen B, Liu S, Gong Y, Zhang L. Combined Phacovitrectomy with Metallic Intraocular Foreign Body Removal through Corneal Incision Using A Novel "Magnetic Conduction" Technique. Retina. 2023;43(12):2157–61.

18. Chen X, Chen Y, Zhong C, Zeng Y, Luo W, Li S. The efficacy and safety of airway foreign body removal by balloon catheter via flexible bronchoscope in children - A retrospective analysis. Int J Pediatr Otorhinolaryngol. 2016;82:88–91.

19. Deng WL, & Li, C. Y. Progress in Diagnosis and Treatment of Tracheobronchial Foreign Body in Adults. Progress in Clinical Medicine. 2022;12(4):2487–92.

20. Hou YT, Wei YH, Liao CK, Lin CF. Personalized multidisciplinary approach of orbital apex foreign body: A case report and literature review. Taiwan J Ophthalmol. 2022;12(3):374–7.

21. Xue Z, Gai Y, Wu Y, liu Z, Li Z. Wearable mechanical and electrochemical sensors for real-time health monitoring. Communications Materials. 2024;5(1):211.

22. Linh VTN, Han S, Koh E, Kim S, Jung HS, Koo J. Advances in wearable electronics for monitoring human organs: Bridging external and internal health assessments. Biomaterials. 2025;314:122865.

23. Yoon H, Dagdeviren C. Towards device technologies non-invasive to our daily lives. Nature Communications. 2025;16(1):1027.

24. Burden of disease scenarios for 204 countries and territories, 2022-2050: a forecasting analysis for the Global Burden of Disease Study 2021. Lancet. 2024;403(10440):2204–56.

25. Zhangming; CHSHWZLLWSQJC. Analysis of the incidence and burden of intraocular foreign bodies in China based on age-period-cohort model. Chinese journal of ophthalmology. 2023;59(08):650–6.

26. Zhang Y, Zhang M, Jiang C, Qiu HY. Intraocular foreign bodies in china: clinical characteristics, prognostic factors, and visual outcomes in 1,421 eyes. Am J Ophthalmol. 2011;152(1):66–73.e1.

27. Rodríguez D, Thirumavalavan N, Pan S, Apoj M, Butaney M, Gross MS, et al. Epidemiology of genitourinary foreign bodies in the united states emergency room setting and its association with mental health disorders. International Journal of Impotence Research. 2019;32(4):426–33.

28. Ulas AB, Aydin Y, Eroglu A. Foreign body aspirations in children and adults. The American Journal of Surgery. 2022;224(4):1168–73.

29. Schuldt T, Großmann W, Weiss NM, Ovari A, Mlynski R, Schraven SP. Aural and nasal foreign bodies in children – Epidemiology and correlation with hyperkinetic disorders, developmental disorders and congenital malformations. International Journal of Pediatric Otorhinolaryngology. 2019;118:165–9.

30. Bakhshaee M, Hebrani P, Shams M, Salehi M, Ghaffari A, Rajati M. Psychological status in children with ear and nose foreign body insertion. International Journal of Pediatric Otorhinolaryngology. 2017;92:103–7.

31. Yuan M, Lu Q. Trends and Disparities in the Incidence of Intraocular Foreign Bodies 1990–2019: A Global Analysis. Frontiers in Public Health. 2022;10.

32. Hoskin AK, Mackey DA, Keay L, Agrawal R, Watson S. Eye Injuries across history and the evolution of eye protection. Acta Ophthalmologica. 2019;97(6):637–43.

33. Javadrashid R, Fouladi DF, Golamian M, Hajalioghli P, Daghighi MH, Shahmorady Z, et al. Visibility of different foreign bodies in the maxillofacial region using plain radiography, CT, MRI and ultrasonography: an in vitro study. Dentomaxillofac Radiol. 2015;44(4):20140229.

34. Kufel J, Bargieł-Łączek K, Koźlik M, Czogalik Ł, Dudek P, Magiera M, et al. Chest X-ray Foreign Objects Detection Using Artificial Intelligence. Journal of Clinical Medicine. 2023;12(18):5841.

35. Soomro N CM, Soomro M, Asif N, Saurman E, Lyle D, Sanders R. Design, Development, and Evaluation of an Injury Surveillance App for Cricket: Protocol and Qualitative Study. JMIR Mhealth Uhealth. 2019;7(1).

